# Comparison of incidence and prognosis of myocardial injury in patients with COVID-19-related respiratory failure and other pulmonary infections: a contemporary cohort study

**DOI:** 10.1101/2023.06.09.23291222

**Authors:** Manoela Astolfi Vivan, Vania Naomi Hirakata, Maria Antônia Torres Arteche, Débora Marques de Araujo, Sandra Cristina Pereira Costa Fuchs, Flávio Danni Fuchs

## Abstract

**Background:** Myocardial injury (MI) is frequent in critically ill patients with COVID-19, but its pathogenesis remains unclear. We hypothesized that MI is not solely due to viral infection by SARS-CoV-2, but rather due to the common pathophysiological mechanisms associated with severe pulmonary infections and respiratory failure.

**Methods:** Contemporary and comparative cohort study designed to compare the incidence of MI in patients with acute respiratory failure caused by COVID-19 to that of patients with other pulmonary infections. In addition, we aim to investigate whether MI is a distinct risk factor for in-hospital mortality in patients with COVID-19-related respiratory failure compared to those with non-COVID-19 infections.

**Results:** The study included 1444 patients with COVID-19 [55.5% men; age 58 (46;68) years] and 182 patients with other pulmonary infections [46.9% men; age 62 (44;73) years]. The incidence of MI at ICU admission was lower in COVID-19 patients (36.4%) compared to non-COVID-19 patients (56%), and this difference persisted after adjusting for age, sex, coronary artery disease, heart failure, SOFA score, lactate, and C-reactive protein [RR 0.84 (95% CI, 0.71-0.99)]. MI at ICU admission was associated with a 59% increase in mortality [RR 1.59 (1.36-1.86); P<0.001], and there was no significant difference in the mortality between patients with COVID-19 and those with other pulmonary infections (P=0.271).

**Conclusion:** Myocardial injury is less frequent in patients with critical COVID-19 pneumonia and respiratory failure compared to those with other types of pneumonia. The occurrence of MI is a significant risk factor for in-hospital mortality, regardless of the etiology of the pulmonary infection.

## INTRODUCTION

The coronavirus disease 2019 (COVID-19) pandemic has resulted in over 500 million SARS-CoV-2 infections globally, with more than 6 million deaths reported. Myocardial injury, defined as an elevated serum troponin level higher than the 99th percentile of a reference population, is a common finding in hospitalized COVID-19 patients [1]. Previous studies have reported the frequency of myocardial injury in COVID-19 patients to range from 9.2 to 63.5% [2,3,4,5,6], with a well-established association with worse outcomes and increased mortality [2,3,4].

Despite multiple proposed mechanisms, including hypoxemia, myocarditis, cytokine storm, systemic inflammation, microvascular dysfunction, vasculitis, and coronary heart disease, the pathogenesis of myocardial injury in COVID-19 patients remains unclear [7]. While some case reports have suggested an association between SARS-CoV-2 infection and myocarditis [8–13], few studies have provided histological confirmation of myocarditis [14–16]. In only one study with two reported cases, histological examination confirmed myocarditis with the identification of viral genome in myocardial cells [14]. The histopathologic heart findings observed during autopsies of COVID-19 non-survivors do not meet the criteria for myocarditis [17].

It is important to note that myocardial injury is not specific to COVID-19 and is frequently observed in critically ill patients due to other causes as well. A systematic review of 20 studies involving 3278 patients reported incidences of myocardial injury ranging from 12% to 85%, with a median of 43% (IQ 21-59%) among intensive care patients [18]. Furthermore, the review demonstrated that elevated troponin was independently associated with an increased risk of death in this population (OR 2.5; 95% confidence interval 1.9 to 3.4; P< 0.001) [18]. Thus, we postulated that myocardial injury observed in COVID-19 patients was not solely due to viral infection by SARS-CoV-2, but rather due to the common pathophysiological mechanisms associated with severe pulmonary infections and respiratory failure.

There is currently no comparative study examining the frequency of myocardial injury in contemporary cohorts of critically ill patients with respiratory failure caused by COVID-19 and those with respiratory failure caused by non-COVID-19 etiologies. Therefore, the primary objective of this study is to compare the incidence of myocardial injury in patients with acute respiratory failure due to COVID-19 with that of patients with respiratory failure caused by other pulmonary infections. It is also unclear if the occurrence of myocardial injury has a distinct influence on the prognosis of patients with pulmonary COVID-19 compared to those with non-COVID-19 infections. Thus, we have addressed this issue as a secondary objective of our study.

## METHODS

We conducted a retrospective contemporary cohort study that included all patients admitted to the intensive care units of Hospital de Clínicas de Porto Alegre (HCPA), a tertiary care university-affiliated hospital, from March 2020 to June 2021, with respiratory failure attributed to pulmonary infection. The HCPA Research Ethics Committee approved the study (number 48398721700005327), and the patient’s informed consent was waived due to the retrospective nature of data collection.

The electronic medical records of all adult patients admitted to the intensive care units of HCPA with respiratory failure attributed to pulmonary infection were reviewed. Acute respiratory failure was defined by the presence of one of the following criteria: PaO2 < 60mm/Hg or SpO2 ≤ 90% with 0.21 FiO2. COVID-19 diagnosis was established based on positive results of nasopharyngeal swabs tested by RT-PCR or antigen testing. All patients included in the study had either RT-PCR or antigen testing for Sars-CoV-2 performed. Patients were either discharged or had died at the time of data collection and analysis.

We collected and recorded clinical data, demographic characteristics, medical history, laboratory tests, and outcomes during hospitalization. Data related to laboratory results and clinical data at ICU admission were considered only if the interval between admission and processing of laboratory data was less than 48 hours. We used a chemiluminescence microparticle immunoassay (Alinity i STAT High Sensitive Troponin-I Reagent Kit, Abbott Laboratories, Lake Forest, IL, USA) for the quantitative determination of cardiac troponin I. For patients who had more than one troponin measurement within 48 hours of admission, we used the highest value recorded.

The primary objective was to determine the proportion of patients with myocardial injury upon ICU admission, as indicated by a high-sensitivity cardiac troponin I value greater than the 99th percentile of a healthy reference population (34.2 pg/mL for men; 15.6 pg/mL for women). The extent of myocardial injury was also evaluated based on the degree of troponin elevation, which was categorized as less than the upper limit of normal (ULN), between 1 and 5 times ULN, between 5 and 10 times ULN, and greater than 10 times ULN, and also assessed as a continuous variable. Patients who had type 1 or type 2 myocardial infarction or did not undergo troponin testing were excluded from the analysis. We compared the association between myocardial injury and in-hospital mortality as well as a composite outcome (in-hospital death, pulmonary embolism, or renal replacement therapy) among patients with respiratory failure due to COVID-19 pneumonia and those with pneumonia caused by non-COVID-19 etiologies, both overall and within each group.

The Statistical Package for the Social Sciences, version 20.0® (Cary, USA) was used to perform statistical analyses. Patients were classified into subgroups based on their COVID-19 diagnosis or other pulmonary infections. The normal distribution of continuous variables was assessed using a histogram and the Shapiro-Wilk test. Descriptive statistics were presented as frequencies (%) for categorical data, means and standard deviations (SD) for continuous data with normal distribution, and median and interquartile range (IQR) for continuous data without normal distribution. Student’s t-test or Mann-Whitney’s test was used for continuous variables, and the chi-square test or Fisher’s exact test was used for categorical variables to compare between groups when appropriate.

A Poisson regression model with robust variance was used to analyze factors associated with myocardial injury, while a Gamma regression model was employed to examine factors associated with troponin as a continuous variable. The linearity of continuous variables was assessed, and the linearity assumption criteria were met. To evaluate the association of myocardial injury with mortality and the composite outcome, Cox proportional hazard models were used, and the proportional hazard assumption was assessed, with the assumption of proportionality criteria being met. Additionally, a Cox proportional hazard model was utilized to evaluate the interaction between COVID-19 and myocardial injury with in-hospital mortality. Confounding variables were selected based on their association with the dependent variable in the univariate analysis (P <0.1) and their presumed causal association with the outcome. Receiver operating characteristic curves were created to assess the ability of high-sensitivity cardiac troponin I to predict in-hospital mortality in patients with COVID-19 or other pulmonary infections. The area under the ROC curves for each group was compared to test for significant differences. Statistical significance was accepted at P < 0.05.

## RESULTS

Out of the 1615 COVID-19 patients admitted to the ICU during the study period, troponin was assessed within 48 hours of admission for 1444 patients (89.4%) who were included in the study. Similarly, troponin was assessed for 182 (90.1%) of the 202 patients admitted to the ICU with other pulmonary infections within 48 hours of admission and included in the study (Figure 1). No significant differences were observed in demographics, comorbidities, or outcomes between patients who had troponin checked and those who did not, as indicated in Supplements 1 and 2. Furthermore, in the sensitivity analysis, including or excluding patients without troponin checked did not alter the comparison of demographics, comorbidities, laboratory and clinical findings at ICU admission or outcomes between COVID-19 and non-COVID-19 patients (Supplement 3).

**Figure 1.**
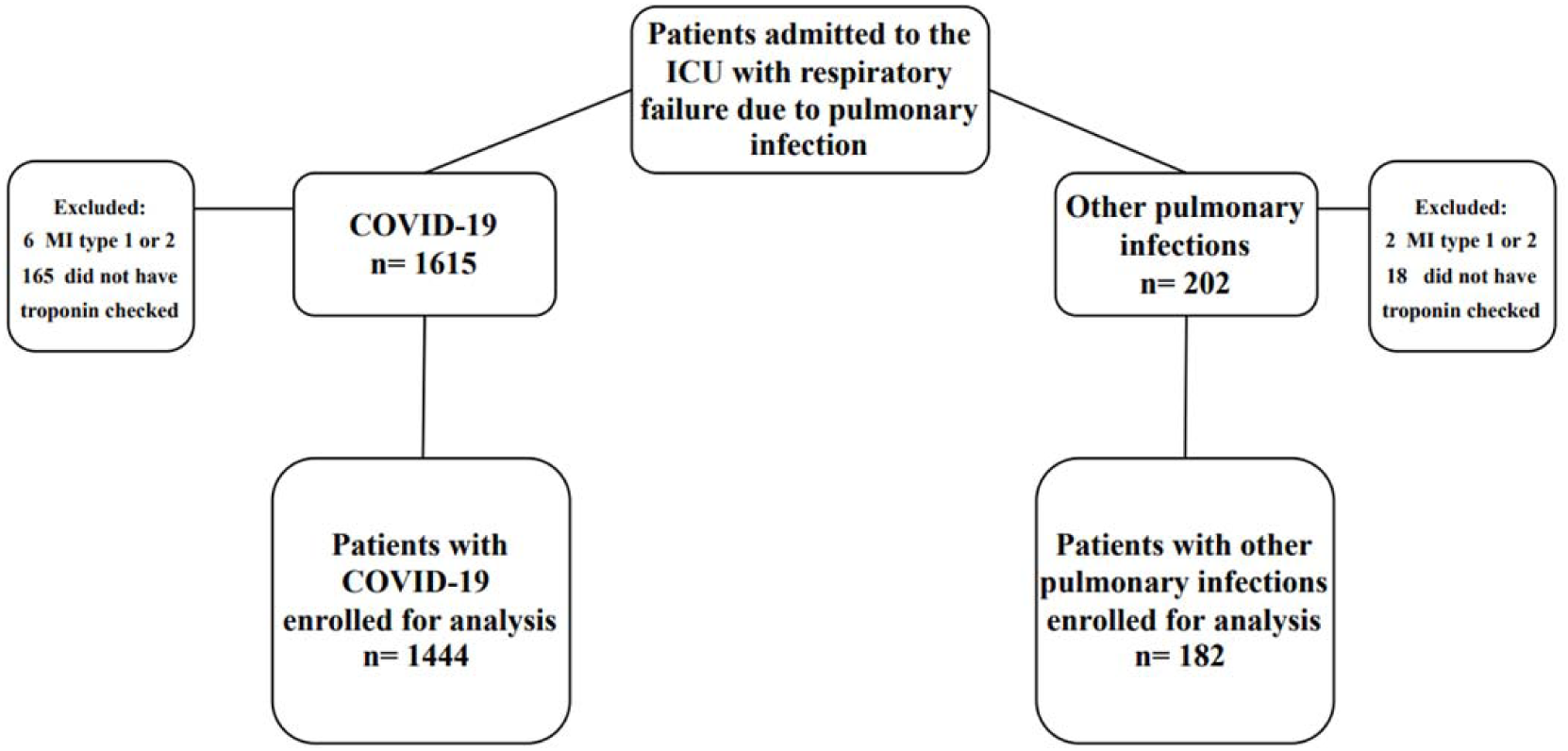
Flow chart of the study. *Abbreviations:* ICU = intensive care unit. MI = myocardial infarction. COVID-19 = coronavirus disease 19

The median age of COVID-19 patients included in the study was 58 years [interquartile range (IQR): 46-68], and 802 patients (55.5%) were male. Among non-COVID-19 patients, the median age was 62 years (IQR: 44-73), and 85 patients (46.7%) were male. Patients admitted due to non-COVID-19 pulmonary infections had a lower body mass index [30.4 (IQR: 26.5-35.7) vs. 26.5 (IQR: 22.3-31.3); P<0.001] and a higher prevalence of comorbidities such as cerebrovascular disease, heart failure, coronary artery disease, chronic lung disease, and chronic HIV infection, as shown in Table 1. Non-COVID-19 patients had higher Sequential Organ Failure Assessment (SOFA) scores, a greater need for invasive mechanical ventilation, and a higher need for vasopressors at ICU admission, as demonstrated in Table 1. Conversely, non-COVID-19 patients had higher PaO2/FiO2 ratios, indicating better gas exchange compared to COVID-19 patients (Table 1).

**Table 1.**
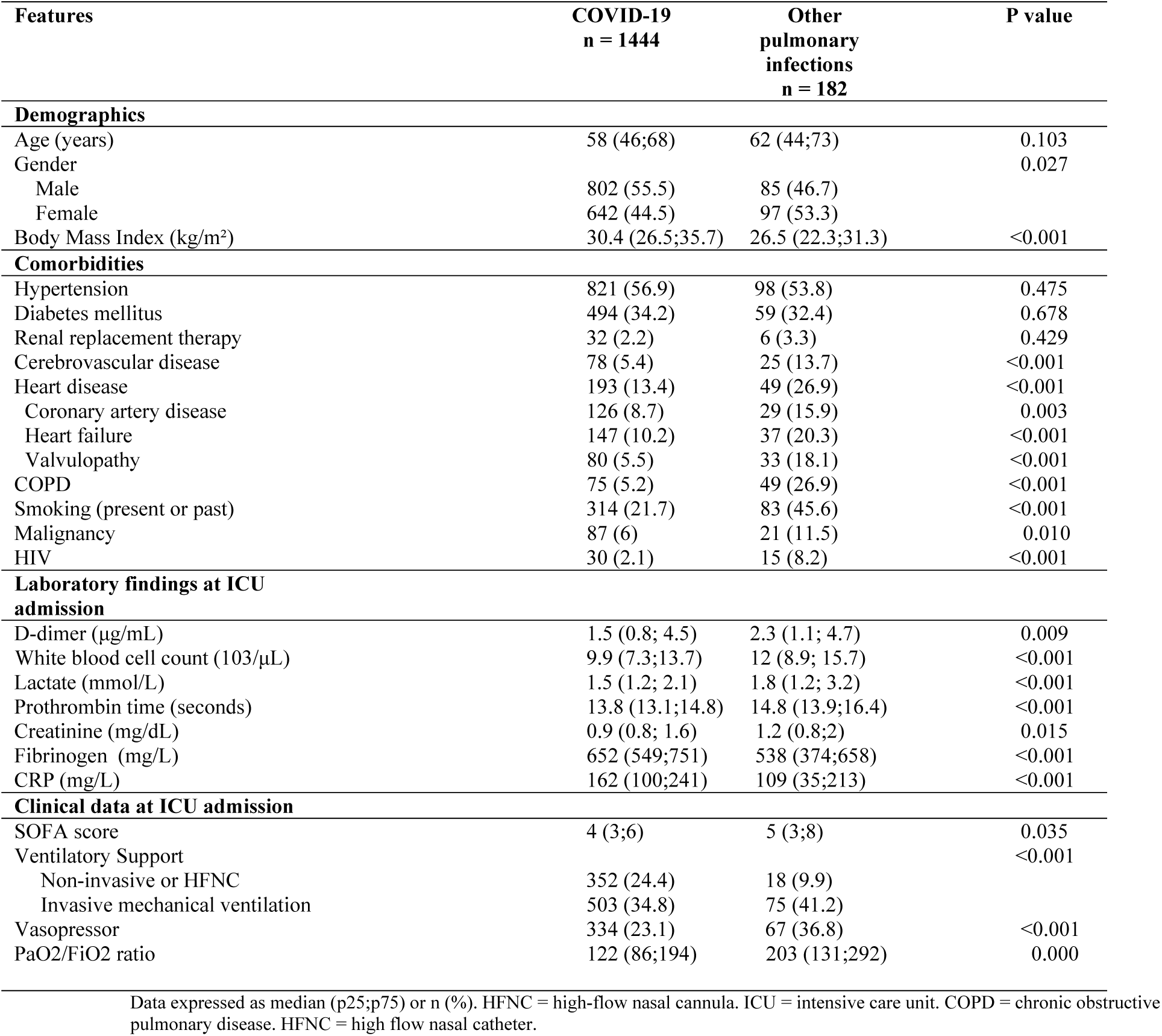
Characteristics of patients admitted to the ICU with respiratory failure attributed to COVID-19 or other pulmonary infections.

| Features | COVID-19<br>n = 1444 | Other<br>pulmonary<br>infections<br>n = 182 | P value |
| --- | --- | --- | --- |
| **Demographics** |  |  |  |
| Age (years) | 58 (46;68) | 62 (44;73) | 0.103 |
| Gender |  |  | 0.027 |
| Male | 802 (55.5) | 85 (46.7) |  |
| Female | 642 (44.5) | 97 (53.3) |  |
| Body Mass Index (kg/m<sup>2</sup>) | 30.4 (26.5;35.7) | 26.5 (22.3;31.3) | <0.001 |
| **Comorbidities** |  |  |  |
| Hypertension | 821 (56.9) | 98 (53.8) | 0.475 |
| Diabetes mellitus | 494 (34.2) | 59 (32.4) | 0.678 |
| Renal replacement therapy | 32 (2.2) | 6 (3.3) | 0.429 |
| Cerebrovascular disease | 78 (5.4) | 25 (13.7) | <0.001 |
| Heart disease | 193 (13.4) | 49 (26.9) | <0.001 |
| Coronary artery disease | 126 (8.7) | 29 (15.9) | 0.003 |
| Heart failure | 147 (10.2) | 37 (20.3) | <0.001 |
| Valvulopathy | 80 (5.5) | 33 (18.1) | <0.001 |
| COPD | 75 (5.2) | 49 (26.9) | <0.001 |
| Smoking (present or past) | 314 (21.7) | 83 (45.6) | <0.001 |
| Malignancy | 87 (6) | 21 (11.5) | 0.010 |
| HIV | 30 (2.1) | 15 (8.2) | <0.001 |
| **Laboratory findings at ICU admission** |  |  |  |
| D-dimer (µg/mL) | 1.5 (0.8; 4.5) | 2.3 (1.1; 4.7) | 0.009 |
| White blood cell count (103/µL) | 9.9 (7.3;13.7) | 12 (8.9; 15.7) | <0.001 |
| Lactate (mmol/L) | 1.5 (1.2; 2.1) | 1.8 (1.2; 3.2) | <0.001 |
| Prothrombin time (seconds) | 13.8 (13.1;14.8) | 14.8 (13.9;16.4) | <0.001 |
| Creatinine (mg/dL) | 0.9 (0.8; 1.6) | 1.2 (0.8;2) | 0.015 |
| Fibrinogen (mg/L) | 652 (549;751) | 538 (374;658) | <0.001 |
| CRP (mg/L) | 162 (100;241) | 109 (35;213) | <0.001 |
| **Clinical data at ICU admission** |  |  |  |
| SOFA score | 4 (3;6) | 5 (3;8) | 0.035 |
| Ventilatory Support |  |  | <0.001 |
| Non-invasive or HFNC | 352 (24.4) | 18 (9.9) |  |
| Invasive mechanical ventilation | 503 (34.8) | 75 (41.2) |  |
| Vasopressor | 334 (23.1) | 67 (36.8) | <0.001 |
| PaO<sub>2</sub>/FiO<sub>2</sub> ratio | 122 (86;194) | 203 (131;292) | 0.000 |

Data expressed as median (p25;p75) or n (%). HFNC = high-flow nasal cannula. ICU = intensive care unit. COPD = chronic obstructive pulmonary disease. HFNC = high flow nasal catheter.

The proportion of patients with myocardial injury at ICU admission was lower among COVID-19 patients (36.4%) compared to non-COVID-19 patients (56%) [Figure 2; relative risk (RR) 0.64; 95% confidence interval (CI) 0.56–0.75)]. Although this association weakened with covariate adjustment, it remained statistically significant after controlling for age, sex, coronary artery disease, heart failure, Sequential Organ Failure Assessment (SOFA) score (creatinine, total bilirubin, PaO2/FiO2 ratio, mean arterial pressure/vasopressor, Glasgow Coma Scale, platelets), lactate, and C-reactive protein [RR 0.84 (95% CI, 0.71-0.99)]. When troponin levels were assessed as a continuous variable, they were also lower in COVID-19 patients compared to non-COVID-19 patients [Table 2; median (interquartile range) 11.6 (9.9-53.7) vs. 35.5 (9.9-218), p <0.001], and this difference remained statistically significant after adjusting in a gamma regression model a gamma regression model for age, sex, coronary artery disease, heart failure, SOFA score (creatinine, total bilirubin, PaO2/FiO2 ratio, mean arterial pressure/vasopressor, Glasgow Coma Scale, platelets), lactate, and C-reactive protein using (P=0.042).

**Figure 2.**
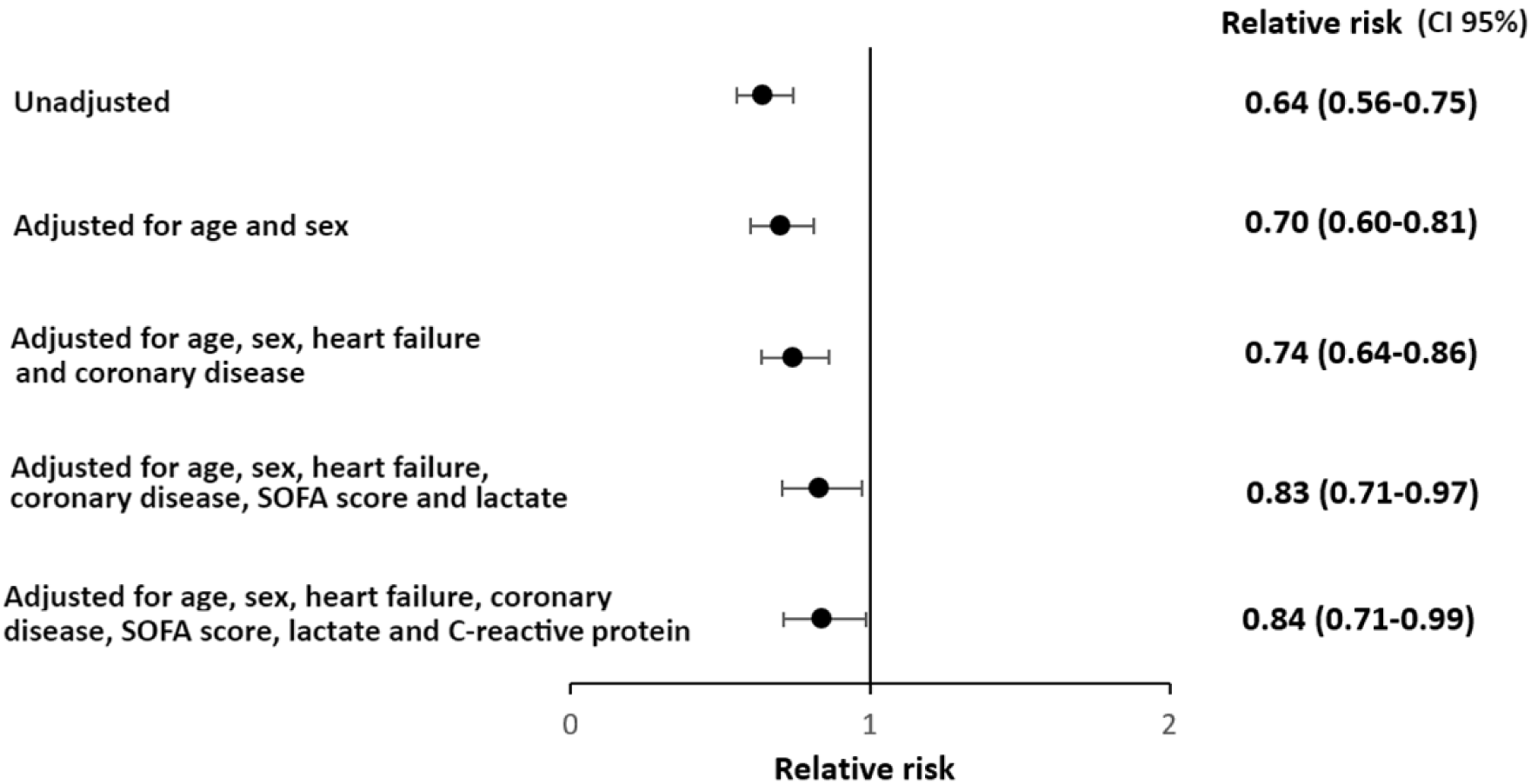
Myocardial injury relative risk (CI 95%) for COVID-19 *vs* non COVID-19 patients. Adjusted by robust Poisson regression model for age, sex, coronary artery disease, heart failure, SOFA score (creatinine, total bilirubin, PaO2/FiO2 ratio, mean arterial pressure/vasopressor, Glasgow Coma Scale, platelets), lactate and C-reactive protein.

**Table 2.**
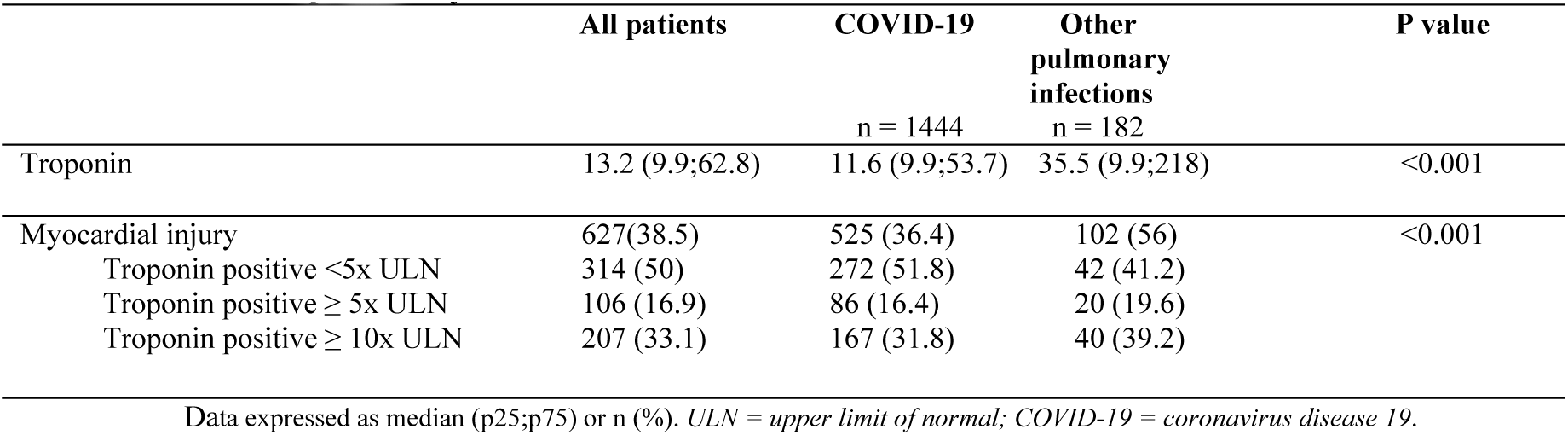
Myocardial injury among patients admitted to the ICU with respiratory failure attributed to COVID-19 or other pulmonary infections.

COVID-19 patients had higher rates of in-hospital death, the composite outcome (in-hospital death, pulmonary embolism or renal replacement therapy), longer hospital stays, longer ICU stays, and longer mechanical ventilation duration compared to non-COVID-19 patients (Table 3; P<0.001). Although pulmonary embolism was more frequently diagnosed among COVID-19 patients (20.6% vs 5.5%), it is worth noting that 822/1615 (50.9%) of these patients underwent a computed tomography pulmonary angiogram (CTPA), while only 47/202 (23.3%) of non-COVID-19 patients underwent a CTPA. The in-hospital mortality rate was 41% among COVID-19 patients and 26.4% among patients with other pulmonary infections.

**Table 3.** Outcomes in patients admitted to the ICU with respiratory failure attributed to COVID-19 or other pulmonary infections.

|  | COVID-19<br>n = 1444 | Other<br>pulmonary<br>infections<br>n = 182 | P value |
| --- | --- | --- | --- |
| **Outcomes** |  |  |  |
| Renal replacement therapy (new) | 339 (23.5) | 21 (11.5) | < 0.001 |
| Pulmonary embolism | 298 (20.6) | 10 (5.5) | <0.001 |
| Non survivor | 592 (41) | 48 (26.4) | <0.001 |
| Composite | 838 (58) | 66 (36.3) | <0.001 |
| Length of hospital stay | 19 (11;32) | 14 (10;22) | <0.001 |
| Length of ICU stay | 10 (6;21) | 4 (1;12) | <0.001 |
| Length of mechanical ventilation | 13 (7;24) | 6 (4;11) | < 0.001 |

Data expressed as median (p25;p75) or n (%). Composite outcome = in-hospital death, pulmonary embolism, renal replacement therapy (new). ICU = intensive care unit.

The mortality rate was significantly higher in COVID-19 patients with troponin levels >5x ULN (49.8%) compared to those with troponin levels under the ULN (26.4%; P<0.001; Figure 3). Similarly, non-COVID-19 patients with higher troponin levels had a higher mortality rate (31.7%) compared to those with troponin levels under the ULN (16.3%; P=0.032; Figure 3).

**Figure 3.**
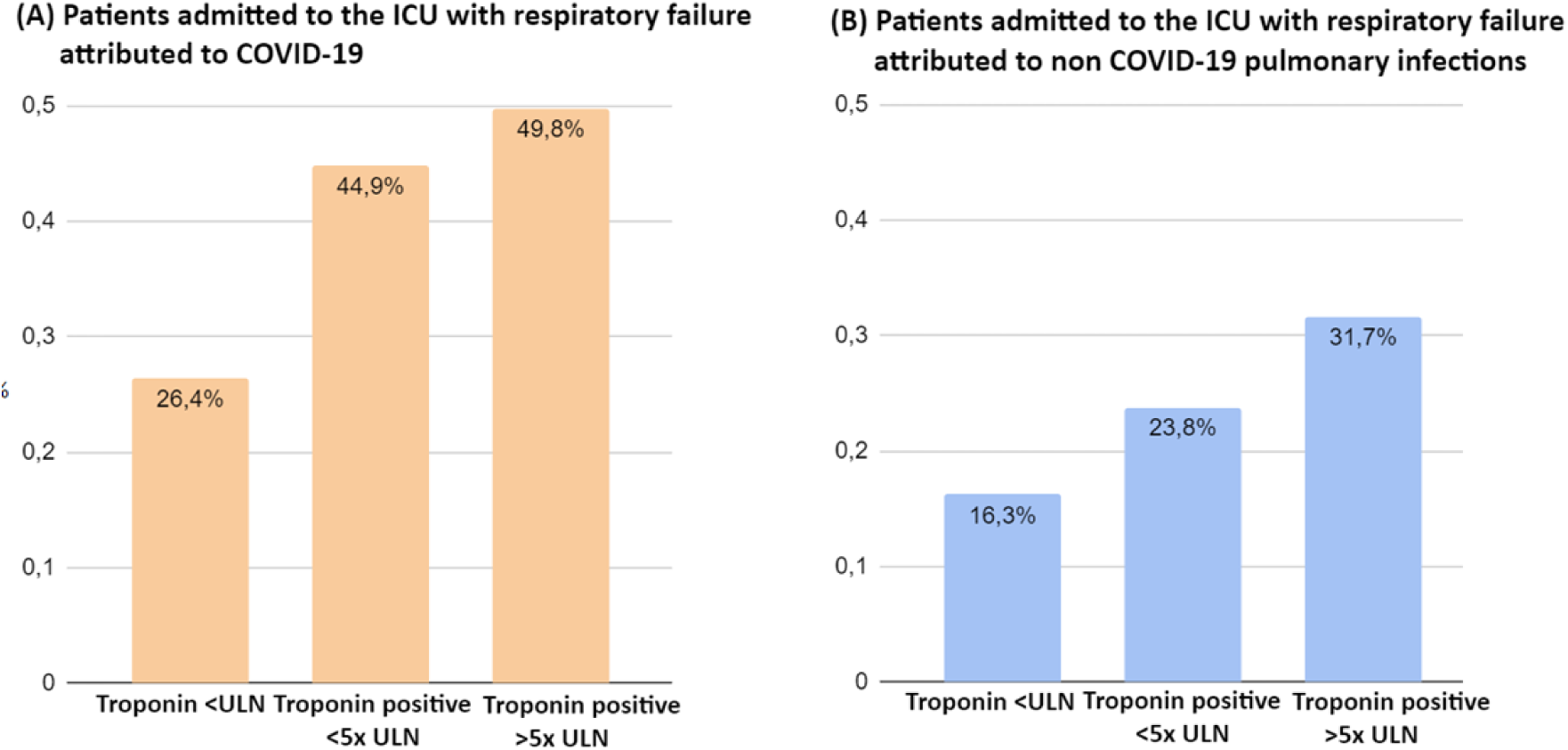
30-day mortality by category of troponin level at ICU admission. (A) Patients admitted to the ICU with respiratory failure attributed to COVID-19. P<0.001 for difference in proportions. (B) Patients admitted to the ICU with respiratory failure attributed to non COVID-19 pulmonary infections. P=0.032 for difference in proportions. *Legend: ULN = upper limit of normal; COVID-19 = coronavírus disease 19*

The presence of myocardial injury at ICU admission was associated with a 59% increase in mortality [RR 1.59 (95% CI, 1.36-1.86), P<0.001]. This association attenuated but remained statistically significant after adjusting for age, sex, and SOFA score [RR 1.21 (95% CI, 1.01-1.44), P=0.034]. The association between myocardial injury and mortality was also present when troponin was evaluated as a continuous variable (P = 0.026). There was no significant interaction effect between COVID-19 and non-COVID-19 infections regarding the association of myocardial injury with in-hospital death (P=0.271). The AUC for high-sensitivity cardiac troponin I to predict in-hospital mortality was 0.66 (95% CI, 0.63-0.69) for COVID-19 patients and 0.63 (95% CI, 0.53-0.72) for other pulmonary infections (Figure 4). There was no statistically significant difference in the C-statistic for the AUC calculated for high-sensitivity cardiac troponin I to predict in-hospital mortality in COVID-19 patients compared to other pulmonary infections (P=0.572).

**Figure 4.**
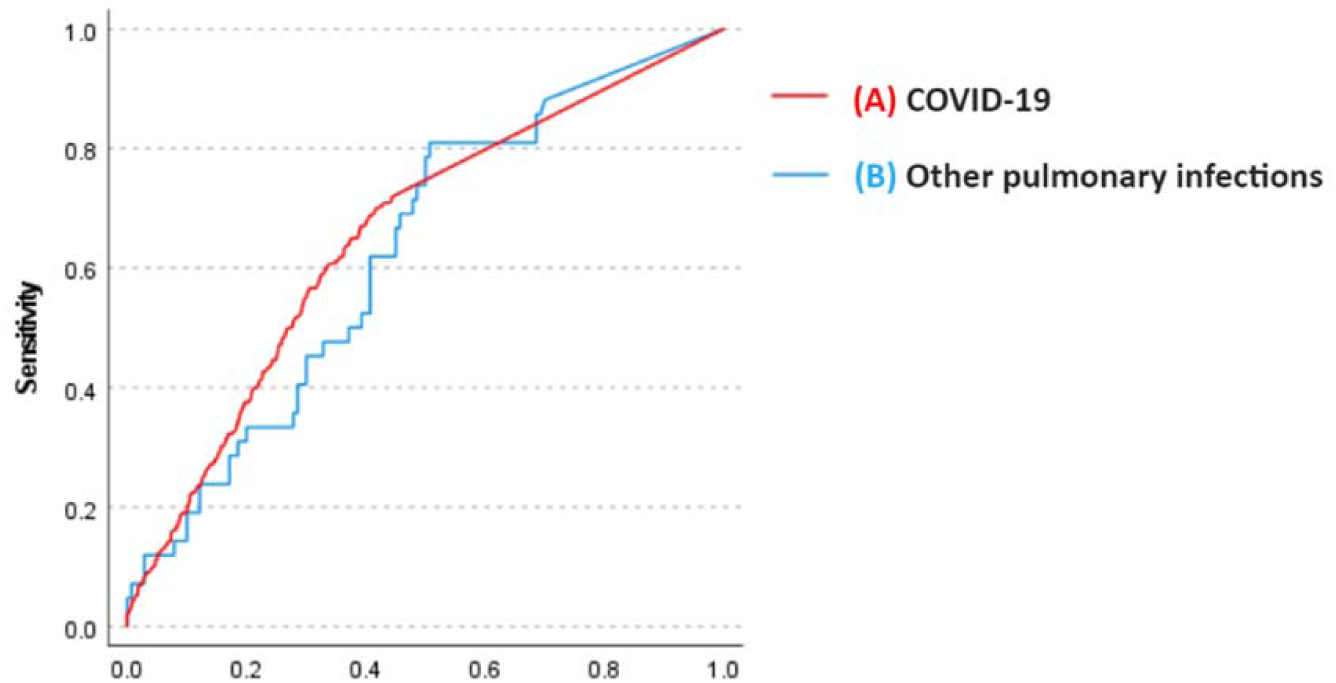
Receiver operating characteristics curves calculated for high-sensitivity cardiac troponin I to predict in-hospital mortality. (A) COVID-19 patients. Area under de curve(AUC) 0,656(95% CI 0.626-0.685). (B) Other pulmonary infections. Area under de curve(AUC) 0,628 (95% CI 0.535-0.720). There was no statistically significant difference in C-statistic for AUC calculated for high-sensitivity cardiac troponin I to predict in-hospital mortality in COVID-19 patients compared to other pulmonary infections (p=0.572).

## DISCUSSION

In this retrospective contemporary cohort study of critically ill patients with respiratory failure, the incidence of myocardial injury was less common in patients with COVID-19 pneumonia than in patients with other pulmonary infections. The occurrence of myocardial injury was a risk factor for in-hospital mortality, regardless of whether the infection was caused by COVID-19 or other agents. These findings highlight the additional risk posed by myocardial injury in patients with severe pneumonia and respiratory failure, and suggest that it is not directly caused by the infectious agent but rather is more likely due to the multisystem organ dysfunction secondary to Severe Acute Respiratory Syndrome (SARS).

During the early stages of the pandemic, an alarming incidence of myocardial injury was detected among critically ill COVID-19 patients [2,3,4,5,6], leading to the elaboration of hypotheses to explain this incidence. Among them, it was proposed that the pulmonary and cardiovascular damage could be mediated by a functional cell entry receptor of Sars-CoV-2, a type 2 Angiotensin Converting Enzyme receptor (ACE2), which is abundantly expressed on the surface of cells in the lungs and cardiovascular system [19]. This hypothesis was based on an apparent higher risk of complications by COVID-19 infection identified in patients taking Angiotensin Receptor Blockers (ARB) [19,20]. The demonstration that a higher risk for severe COVID-19 infection was not influenced by the use of ARB, suggesting that first observations were confounded by hypertension, turned this hypothesis unlikely [21,22]. The hypothesis that myocardial injury could be caused by myocarditis by COVID-19 was also unlikely. The genome of the virus has been identified in the myocardium in a few studies [13], and the histopathologic findings observed described in autopsies of COVID-19 non-survivors are not suggestive of myocarditis. [7]. Myocardial injury seen in patients with severe COVID-19 infection, particularly with severe pneumonia and respiratory failure, could be secondary to unspecific supply-demand mismatch, cytokine storm, systemic inflammation, microvascular dysfunction, vasculitis, and coronary thrombosis, described in patents with Acute Respiratory Distress Syndrome [23]. Our findings are in accordance with this hypothesis.

To date, only two studies have compared the incidence of myocardial injury in cohorts of patients with respiratory failure caused by Sars-CoV-2 and other agents [5,6]. However, these studies had limitations, including smaller sample sizes of patients with COVID-19 and the use of historical controls. In the study conducted at the hospitals of the Johns Hopkins Healthcare System, the incidence of myocardial injury was found to be similar (around 50%) in patients with COVID-19 and non-COVID-19 pneumonia [5]. The 2-fold increased hazard for mortality was no longer statistically significant after adjusting for covariates. The study conducted in Austria and Germany found higher incidences of myocardial injury and a higher incidence in patients with other types of pneumonia (96.4%) compared to those with COVID-19 (78.1%) [6], but did not report the association of myocardial injury with the risk of mortality.

Non-comparative studies that included only patients with respiratory failure due to Sars-CoV-2 found an incidence of myocardial injury that was similar to that observed in our study [2,3,4]. Similarly, cohorts of patients with respiratory failure caused by other infectious agents, as well as those with critical illness from other causes, have shown an incidence of myocardial injury that is approximately similar to our findings [18,24,25].

Our study has limitations, mainly related to retrospective data collection. Nonetheless, the criteria for the selection of participants with and without COVID-19 respiratory failure were similar, and the cohorts were contemporary and managed in an ICU with equal resources and medical expertise. Although not all patients had myocardial injury assessed in the first 48 hours from ICU admission (10% of missing troponin I US at ICU admission), the baseline characteristics and outcomes of the patients who did and did not have troponin checked were comparable. Additionally, myocardial injury was diagnosed solely by cardiac markers, without including further cardiac evaluation tests like echocardiography, magnetic resonance imaging, or biopsy. Nevertheless, it is unlikely that the groups differed regarding the findings of these examinations. The strengths of our study include the comparison of contemporary cohorts, the thorough control for a comprehensive set of potential confounders, and the relatively large sample size.

In conclusion, our study provides evidence that myocardial injury is less common in patients with COVID-19 pneumonia and respiratory failure compared to those with other severe pulmonary infections. This finding supports the hypothesis that the occurrence of myocardial injury is secondary to pathophysiological mechanisms associated with serious pulmonary infection and respiratory failure. Additionally, our study found that the presence of myocardial injury is a risk factor for in-hospital mortality, irrespective of the etiology of the pulmonary infection.

The practical implication of these findings is that the key to reducing the risk of myocardial injury and its consequences may be to institute adequate intensive care and support to optimize organ dysfunction.

## Data Availability

The data that support the findings of this study are available from the corresponding author, MAV, upon reasonable request.

## Data Access, Responsibility, and Analysis Statement

MAV had full access to all the data in the study and takes responsibility for the integrity of the data and the accuracy of the data analysis.

## Ethical Approval Statement

All procedures performed in this study were in accordance with the ethical standards of the HCPA Research Ethics Committee and with the Helsinki declaration and its later amendments.

## Informed consent Statement

Not applicable

## Author Contribution Statement

MAV, FF and SF: conceptualization, methodology, investigation, formal analysis, writing. MATA and DMA: investigation and critical review. VNH: statistical analysis and critical review. All the authors reviewed and approved the final manuscript.

---

**Supplement 1.**
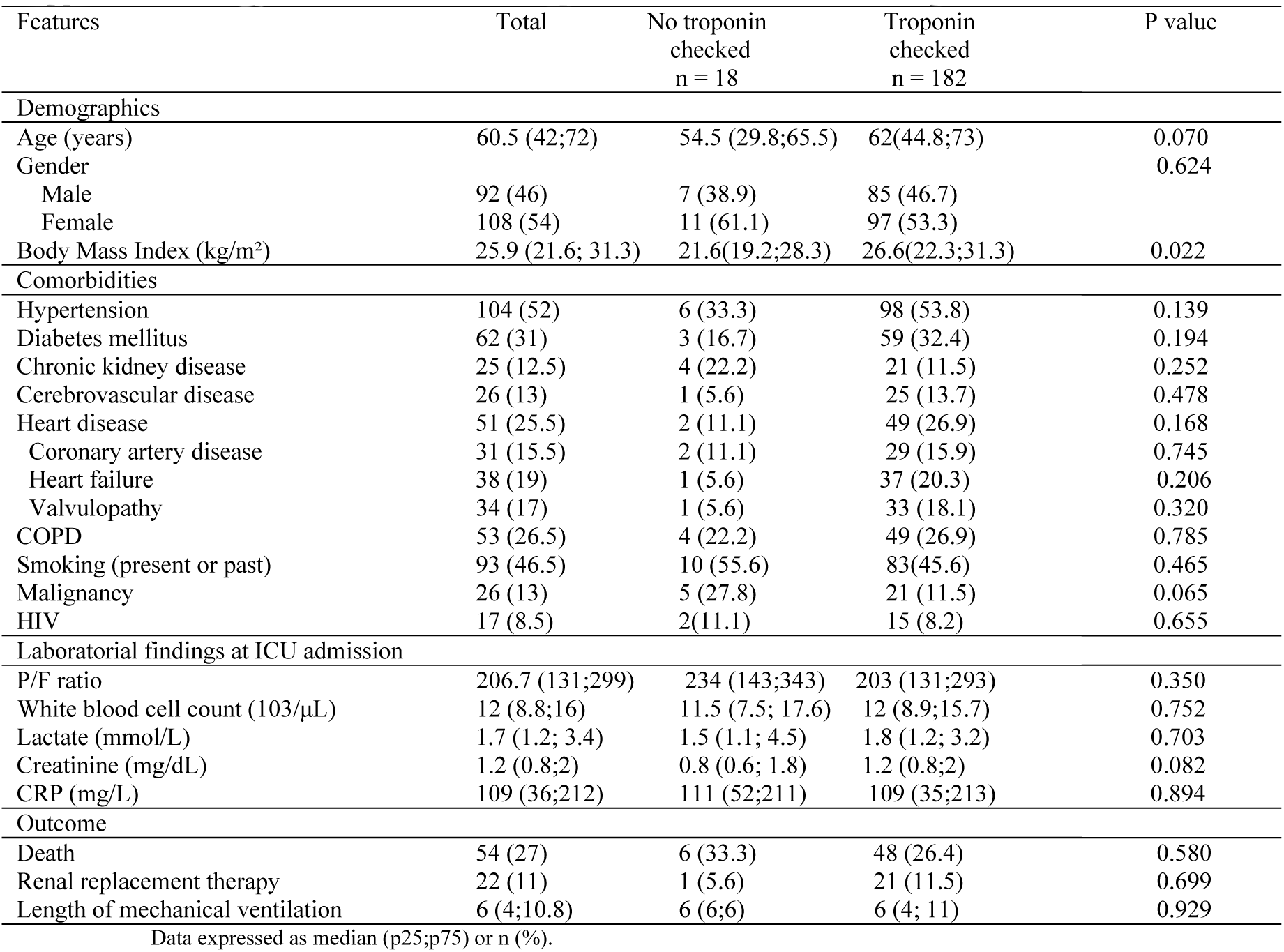
Comparison of non-COVID-19 patients with versus without troponin checked.

| Features | Total | No troponin checked<br>n = 18 | Troponin checked<br>n = 182 | P value |
| --- | --- | --- | --- | --- |
| **Demographics** |  |  |  |  |
| Age (years) | 60.5 (42;72) | 54.5 (29.8;65.5) | 62(44.8;73) | 0.070 |
| Gender |  |  |  | 0.624 |
| Male | 92 (46) | 7 (38.9) | 85 (46.7) |  |
| Female | 108 (54) | 11 (61.1) | 97 (53.3) |  |
| Body Mass Index (kg/m<sup>2</sup>) | 25.9 (21.6; 31.3) | 21.6(19.2;28.3) | 26.6(22.3;31.3) | 0.022 |
| **Comorbidities** |  |  |  |  |
| Hypertension | 104 (52) | 6 (33.3) | 98 (53.8) | 0.139 |
| Diabetes mellitus | 62 (31) | 3 (16.7) | 59 (32.4) | 0.194 |
| Chronic kidney disease | 25 (12.5) | 4 (22.2) | 21 (11.5) | 0.252 |
| Cerebrovascular disease | 26 (13) | 1 (5.6) | 25 (13.7) | 0.478 |
| Heart disease | 51 (25.5) | 2 (11.1) | 49 (26.9) | 0.168 |
| Coronary artery disease | 31 (15.5) | 2 (11.1) | 29 (15.9) | 0.745 |
| Heart failure | 38 (19) | 1 (5.6) | 37 (20.3) | 0.206 |
| Valvulopathy | 34 (17) | 1 (5.6) | 33 (18.1) | 0.320 |
| COPD | 53 (26.5) | 4 (22.2) | 49 (26.9) | 0.785 |
| Smoking (present or past) | 93 (46.5) | 10 (55.6) | 83(45.6) | 0.465 |
| Malignancy | 26 (13) | 5 (27.8) | 21 (11.5) | 0.065 |
| HIV | 17 (8.5) | 2(11.1) | 15 (8.2) | 0.655 |
| **Laboratorial findings at ICU admission** |  |  |  |  |
| P/F ratio | 206.7 (131;299) | 234 (143;343) | 203 (131;293) | 0.350 |
| White blood cell count (103/μL) | 12 (8.8;16) | 11.5 (7.5; 17.6) | 12 (8.9;15.7) | 0.752 |
| Lactate (mmol/L) | 1.7 (1.2; 3.4) | 1.5 (1.1; 4.5) | 1.8 (1.2; 3.2) | 0.703 |
| Creatinine (mg/dL) | 1.2 (0.8;2) | 0.8 (0.6; 1.8) | 1.2 (0.8;2) | 0.082 |
| CRP (mg/L) | 109 (36;212) | 111 (52;211) | 109 (35;213) | 0.894 |
| **Outcome** |  |  |  |  |
| Death | 54 (27) | 6 (33.3) | 48 (26.4) | 0.580 |
| Renal replacement therapy | 22 (11) | 1 (5.6) | 21 (11.5) | 0.699 |
| Length of mechanical ventilation | 6 (4;10.8) | 6 (6;6) | 6 (4; 11) | 0.929 |

Data expressed as median (p25;p75) or n (%).

**Supplement 2.**
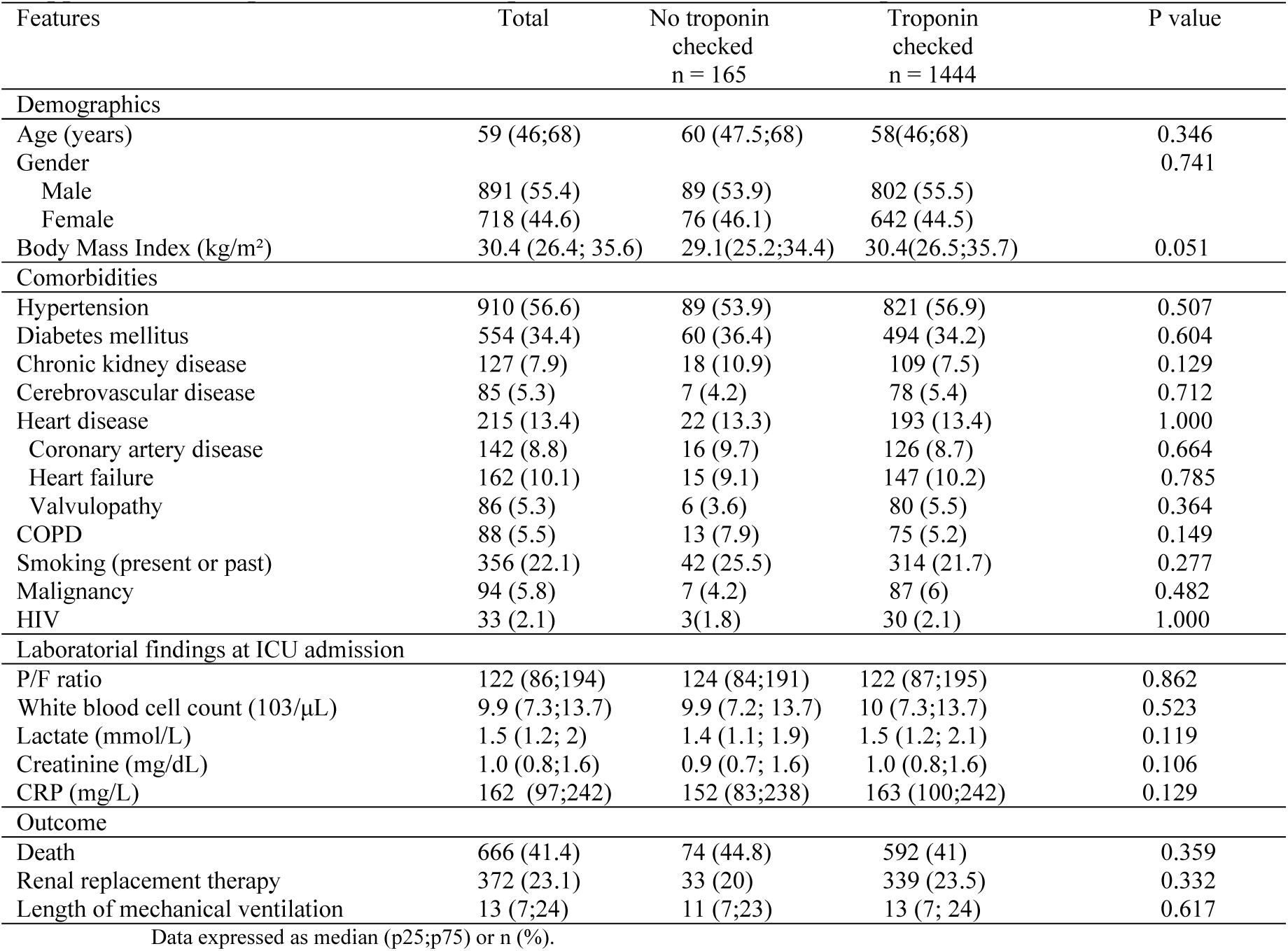
Comparison of COVID-19 patients with versus without troponin checked.

**Supplement 3.** Characteristics of patients admitted to the ICU with respiratory failure attributed to COVID-19 or other pulmonary infections (sensitivity analysis including patients without troponin checked)

| Features | COVID-19<br>n = 1609 | Other<br>pulmonary<br>infections<br>n = 200 | P value |
| --- | --- | --- | --- |
| **Demographics** |  |  |  |
| Age (years) | 59 (46;68) | 60.5 (42;72) | 0.315 |
| Gender |  |  | 0.013 |
| Male | 891 (55.4) | 92 (46) |  |
| Female | 718 (44.6) | 108 (54) |  |
| Body Mass Index (kg/m<sup>2</sup>) | 30.4 (26.4;35.6) | 25.9 (21.6;31.2) | <0.001 |
| **Comorbidities** |  |  |  |
| Hypertension | 910 (56.6) | 104 (52) | 0.227 |
| Diabetes mellitus | 554 (34.4) | 62 (31) | 0.344 |
| Renal replacement therapy | 36 (2.2) | 9 (4.5) | 0.085 |
| Cerebrovascular disease | 85 (5.3) | 26 (13) | <0.001 |
| Heart disease | 215 (13.4) | 51 (25.5) | <0.001 |
| Coronary artery disease | 142 (8.8) | 31 (15.5) | 0.005 |
| Heart failure | 162 (10.1) | 38 (19) | <0.001 |
| Valvulopathy | 86 (5.3) | 34 (17) | <0.001 |
| COPD | 88 (5.5) | 53 (26.5) | <0.001 |
| Smoking (present or past) | 356 (22.1) | 93 (46.5) | <0.001 |
| Malignancy | 94 (5.8) | 26 (13) | <0.001 |
| HIV | 33(2.1) | 17 (8.5) | <0.001 |
| **Laboratory findings at ICU admission** |  |  |  |
| D-dimer (µg/mL) | 1.5 (0.8; 4.5) | 2.4 (1; 5) | 0.012 |
| White blood cell count (103/µL) | 9.9 (7.3;13.7) | 12 (8.8; 15.9) | <0.001 |
| Lactate (mmol/L) | 1.5 (1.2; 2) | 1.7 (1.2; 3.4) | <0.001 |
| Prothrombin time (seconds) | 13.8 (13.1;14.9) | 15 (13.8;16.7) | <0.001 |
| Creatinine (mg/dL) | 0.9 (0.8; 1.6) | 1.2 (0.8;2) | 0.045 |
| Fibrinogen (mg/L) | 648 (546;748) | 541 (375;656) | <0.001 |
| CRP (mg/L) | 162 (97;242) | 109 (36;213) | <0.001 |
| **Clinical data at ICU admission** |  |  |  |
| SOFA score | 4 (3;6) | 5 (3;8) | 0.039 |
| Ventilatory Support |  |  | <0.001 |
| Non-invasive or HFNC | 402 (25) | 21 (10.5) |  |
| Invasive mechanical ventilation | 534 (33.2) | 83 (41.5) |  |
| Vasopressor | 357 (22.2) | 75 (37.5) | <0.001 |
| PaO2/FiO2 ratio | 122 (86;193) | 206 (131;298) | 0.000 |
| **Outcomes** |  |  |  |
| Renal replacement therapy (new) | 372 (23.1) | 22 (11) | < 0.001 |
| Pulmonary embolism | 334 (20.8) | 10 (5) | <0.001 |
| Non survivor | 666 (41.4) | 54 (27) | <0.001 |
| Composite | 930 (57.8) | 72 (36) | <0.001 |
| Length of hospital stay | 19 (11;31) | 13 (9;22) | <0.001 |
| Length of ICU stay | 10 (5;20) | 4 (1;11) | <0.001 |
| Length of mechanical ventilation | 13 (7;24) | 6 (4;10.8) | < 0.001 |

## References

[1] Thygesen K, Alpert JS, Jaffe AS, Chaitman BR, Bax JJ, Morrow DA, White HD; Executive Group on behalf of the Joint European Society of Cardiology (ESC)/American College of Cardiology (ACC)/American Heart Association (AHA)/World Heart Federation (WHF) Task Force for the Universal Definition of Myocardial Infarction. Fourth Universal Definition of Myocardial Infarction (2018). Circulation. 2018 Nov 13;138(20):e618–e651. doi: 10.1161/CIR.0000000000000617. Erratum in: Circulation. 2018 Nov 13;138(20):e652. PMID: 30571511.

[2] Li Y, Pei H, Zhou C, Lou Y. Myocardial Injury Predicts Risk of Short-Term All-Cause Mortality in Patients With COVID-19: A Dose-Response Meta-Analysis. Front Cardiovasc Med. 2022 May 2;9:850447. doi: 10.3389/fcvm.2022.850447. PMID: 35586652; PMCID: PMC9108210.

[3] Changal K, Veria S, Mack S, Paternite D, Sheikh SA, Patel M, Mir T, Sheikh M, Ramanathan PK. Myocardial injury in hospitalized COVID-19 patients: a retrospective study, systematic review, and meta-analysis. BMC Cardiovasc Disord. 2021 Dec 31;21(1):626. doi: 10.1186/s12872-021-02450-3. PMID: 34972516; PMCID: PMC8719604.

[4] Malik P, Patel U, Patel NH, Somi S, Singh J. Elevated cardiac troponin I as a predictor of outcomes in COVID-19 hospitalizations: a meta-analysis. Infez Med. 2020 Dec 1;28(4):500–506. PMID: 33257623.

[5] Metkus TS, Sokoll LJ, Barth AS, Czarny MJ, Hays AG, Lowenstein CJ, Michos ED, Nolley EP, Post WS, Resar JR, Thiemann DR, Trost JC, Hasan RK. Myocardial Injury in Severe COVID-19 Compared With Non-COVID-19 Acute Respiratory Distress Syndrome. Circulation. 2021 Feb 9;143(6):553–565. doi: 10.1161/CIRCULATIONAHA.120.050543. Epub 2020 Nov 13. PMID: 33186055; PMCID: PMC7864609.

[6] Jirak P, Larbig R, Shomanova Z, Fröb EJ, Dankl D, Torgersen C, Frank N, Mahringer M, Butkiene D, Haake H, Salzer HJF, Tschoellitsch T, Lichtenauer M, Egle A, Lamprecht B, Reinecke H, Hoppe UC, Pistulli R, Motloch LJ. Myocardial injury in severe COVID-19 is similar to pneumonias of other origin: results from a multicentre study. ESC Heart Fail. 2021 Feb;8(1):37–46. doi: 10.1002/ehf2.13136. Epub 2020 Dec 17. PMID: 33350605; PMCID: PMC7835505.

[7] Sewanan LR, Clerkin KJ, Tucker NR, Tsai EJ. How Does COVID-19 Affect the Heart? Curr Cardiol Rep. 2023 Mar;25(3):171–184. doi: 10.1007/s11886-023-01841-6. Epub 2023 Mar 10. PMID: 36897483; PMCID: PMC9999058.

[8] Inciardi RM, Lupi L, Zaccone G, et al. Cardiac Involvement in a Patient With Coronavirus Disease 2019 (COVID-19). JAMA Cardiol 2020; 5:819.

[9] Zeng JH, Liu YX, Yuan J, et al. First case of COVID-19 complicated with fulminant myocarditis: a case report and insights. Infection 2020; 48:773.

[10] Hu H, Ma F, Wei X, Fang Y. Coronavirus fulminant myocarditis treated with glucocorticoid and human immunoglobulin. Eur Heart J 2021; 42:206.

[11] Fried JA, Ramasubbu K, Bhatt R, et al. The Variety of Cardiovascular Presentations of COVID-19. Circulation 2020; 141:1930.

[12] Tavazzi G, Pellegrini C, Maurelli M, et al. Myocardial localization of coronavirus in COVID-19 cardiogenic shock. Eur J Heart Fail 2020; 22:911.

[13] Kim IC, Kim JY, Kim HA, Han S. COVID-19-related myocarditis in a 21-year-old female patient. Eur Heart J 2020; 41:1859.

[14] Escher F, Pietsch H, Aleshcheva G, et al. Detection of viral SARS-CoV-2 genomes and histopathological changes in endomyocardial biopsies. ESC Heart Fail 2020; 7:2440.

[15] Salah HM, Mehta JL. Takotsubo cardiomyopathy and COVID-19 infection. Eur Heart J Cardiovasc Imaging 2020; 21:1299.

[16] Nicol M, Cacoub L, Baudet M, et al. Delayed acute myocarditis and COVID-19-related multisystem inflammatory syndrome. ESC Heart Fail 2020.

[17] Fox SE, Akmatbekov A, Harbert JL, Li G, Quincy Brown J, Vander Heide RS. Pulmonary and cardiac pathology in African American patients with COVID-19: an autopsy series from New Orleans. Lancet Respir Med. 2020 Jul;8(7):681–686. doi: 10.1016/S2213-2600(20)30243-5. Epub 2020 May 27. PMID: 32473124; PMCID: PMC7255143.

[18] Lim W, Qushmaq I, Devereaux PJ, Heels-Ansdell D, Lauzier F, Ismaila AS, Crowther MA, Cook DJ. Elevated cardiac troponin measurements in critically ill patients. Arch Intern Med. 2006 Dec 11-25;166(22):2446–54. doi: 10.1001/archinte.166.22.2446. PMID: 17159009.

[19] Hoffmann M, Kleine-Weber H, Schroeder S, Krüger N, Herrler T, Erichsen S, Schiergens TS, Herrler G, Wu NH, Nitsche A, Müller MA, Drosten C, Pöhlmann S. SARS-CoV-2 Cell Entry Depends on ACE2 and TMPRSS2 and Is Blocked by a Clinically Proven Protease Inhibitor. Cell. 2020 Apr 16;181(2):271–280.e8. doi: 10.1016/j.cell.2020.02.052. Epub 2020 Mar 5. PMID: 32142651; PMCID: PMC7102627.

[20] Fang L, Karakiulakis G, Roth M. Are patients with hypertension and diabetes mellitus at increased risk for COVID-19 infection? Lancet Respir Med. 2020;8(4):e21. https://doi.org/10.1016/S2213-2600(20)30116-8

[21] Ferrari F, Martins VM, Fuchs FD, Stein R. Renin-Angiotensin-Aldosterone System Inhibitors in COVID-19: A Review. Clinics (Sao Paulo). 2021 Apr 9;76:e2342. doi: 10.6061/clinics/2021/e2342. PMID: 33852652; PMCID: PMC8009081.

[22] Lopes RD, Macedo AVS, de Barros E Silva PGM, Moll-Bernardes RJ, Dos Santos TM, Mazza L, Feldman A, D’Andréa Saba Arruda G, de Albuquerque DC, Camiletti AS, de Sousa AS, de Paula TC, Giusti KGD, Domiciano RAM, Noya-Rabelo MM, Hamilton AM, Loures VA, Dionísio RM, Furquim TAB, De Luca FA, Dos Santos Sousa ÍB, Bandeira BS, Zukowski CN, de Oliveira RGG, Ribeiro NB, de Moraes JL, Petriz JLF, Pimentel AM, Miranda JS, de Jesus Abufaiad BE, Gibson CM, Granger CB, Alexander JH, de Souza OF; BRACE CORONA Investigators. Effect of Discontinuing vs Continuing Angiotensin-Converting Enzyme Inhibitors and Angiotensin II Receptor Blockers on Days Alive and Out of the Hospital in Patients Admitted With COVID-19: A Randomized Clinical Trial. JAMA. 2021 Jan 19;325(3):254–264. doi: 10.1001/jama.2020.25864. PMID: 33464336; PMCID: PMC7816106.

[23] Metkus TS, Guallar E, Sokoll L, Morrow D, Tomaselli G, Brower R, Schulman S, Korley FK. Prevalence and Prognostic Association of Circulating Troponin in the Acute Respiratory Distress Syndrome. Crit Care Med. 2017 Oct;45(10):1709–1717. doi: 10.1097/CCM.0000000000002641. PMID: 28777195; PMCID: PMC5600678.

[24] Efros O, Soffer S, Leibowitz A, Fardman A, Klempfner R, Meisel E, Grossman E. Risk factors and mortality in patients with pneumonia and elevated troponin levels. Sci Rep. 2020 Dec 10;10(1):21619. doi: 10.1038/s41598-020-78287-1. PMID: 33303788; PMCID: PMC7729902.

[25] Vestjens SMT, Spoorenberg SMC, Rijkers GT, Grutters JC, Ten Berg JM, Noordzij PG, Van de Garde EMW, Bos WJW; Ovidius Study Group. High-sensitivity cardiac troponin T predicts mortality after hospitalization for community-acquired pneumonia. Respirology. 2017 Jul;22(5):1000–1006. doi: 10.1111/resp.12996. Epub 2017 Feb 21. PMID: 28221010.

